# Investigating the association between oral health behaviours and risk behaviours of university students – a quantitative study utilising online questionnaires

**DOI:** 10.1101/2024.08.08.24311694

**Authors:** Tanzeelah Azam, George Kitsaras, Juliana Gomez, Michaela Goodwin

**Author notes:** Corresponding author: Tanzeelah Azam. These authors contributed equally to this work. These authors also contributed equally to this work (George Kitsaras, Juliana Gomez, Michaela Goodwin).

## Abstract

**Background:** Young adults are exposed to a variety of risk-related behaviours such as alcohol, smoking, and changes in dietary habits, which may result in unknown outcomes in their oral health. There is limited evidence on whether different risk behaviours are associated with oral health behaviours in the university student population. This study gathers data on the behaviours of students in their first year of university, which will inform the future development of oral health behaviour change interventions for this population.

**Method:** This longitudinal quantitative survey involved 205 first-year university students, aged 18-24. They completed questionnaires at baseline and a 6-month follow-up interval providing information regarding self-reported oral health status, hygiene routines, risk behaviours (e.g., diet, smoking, alcohol) and attitudes towards digital health.

**Results:** The findings show associations between oral health behaviours with risk behaviours including links with oral care routines, bleeding gums, brushing frequency, with exercise, vaping, and unhealthy food and drink intake. Significant changes over the two-time points were also observed, such as the worsening of the self-reported condition of the teeth (p<0.001), reduction in the self-reported condition of the gums (p=0.004), reduced brushing frequency (p=0.003), less regular dental visits (p=0.013), more students intending to visit their previous dentist rather than finding a new dentist at university (p=0.026), and increased consumption of unhealthy non-alcoholic drinks (p=0.003). Positive changes over time include reduced alcohol consumption frequency and units (p=0.030 and p=0.001), fewer instances of binge drinking (p=0.014), and less frequent consumption of unhealthy foods (p=0.034).

**Conclusion:** The findings highlight the complex relationship between oral heal and risk behaviours in this demographic. Poorer oral health behaviours are linked to engagement in risk behaviours. Therefore, oral health behaviours should be targeted alongside other risk behaviours. Tailored interventions should be developed to improve oral health and behaviour among university students.

## Introduction

Oral health plays a significant role in overall general health and wellbeing [1]. Poor oral health is also associated with many diseases such as heart disease, stroke [2], atherosclerosis [3], diabetes [4], and cognitive decline over time [5]. Poor oral health practices can lead to oral diseases, such as dental caries and gum disease which result in pain and discomfort [6], impact an individual’s ability to consume foods, communicate with others, and influence their overall state of well-being [7, 8]. Oral diseases can also negatively affect the quality of life [9], affect academic performance [10] and lead to time off university for treatment and pain [11]. It is therefore crucial to ensure that the oral health of students is understood and addressed.

Young adults in university are exposed to new experiences during their time of transition from teenagers to adults [12]; this exposure may lead to changes in lifestyle choices which involve engaging in risk-related behaviours such as alcohol consumption, smoking, poor eating habits, poor diet, and lack of exercise [13]. These behaviours adopted during young adulthood can persist into later stages in life [14], potentially leading to future health issues [14, 15]. Moreover, while evidence exists regarding the dental health of young adults, the university setting introduces a period of newfound independence and exposure to various risk behaviours [16]. These behaviours can negatively affect individuals [13]. Consequently, the overall habits of university students may change, potentially exacerbating existing habits and behaviours. Recognising the tendency of university students to adopt unhealthy behaviours is important [17, 18], as it becomes more difficult to promote behaviour change as individuals get older [19].

This area of research remains underexplored with limited evidence available to comprehensively understand the correlation between oral health behaviours and risk behaviours in this demographic, particularly during the key life transition of leaving a family home for the first time and gaining greater independence. Some evidence suggests that individuals in this population are exposed to risk behaviours during this period, however, there is a need for further investigation of the links between these risk behaviours and oral health, observing behaviours that occur over the first year of this life event, and the overall interplay with oral hygiene routines. In our initial patient and public involvement work, we confirmed that university students view oral health as important and are open to participating in oral health research which requires them to share insights regarding their oral hygiene and oral health risk-related behaviours. Overall, this study addresses evidence gaps and aims to build evidence on the profiles of this population with regards to oral health and risk-related behaviours. This study successfully achieved its aim by providing valuable insights and identifying areas for targeted intervention. Developing an oral health intervention tailored to this demographic would require a deeper understanding of the links to facilitate positive behaviour change.

## Aims and objectives

This study aimed to comprehensively assess data regarding self-reported oral health, dental attendance, oral hygiene behaviours, and health-related risk behaviours among university students, aiming to longitudinally track and assess changes or stability in the behaviours within the same group of university students who have recently left home. This aim will be achieved through the following objectives: employ a quantitative survey in a university student population to gather self-reported data describing: (a) oral health status and dental attendance, (b) oral hygiene routine and behaviours and (c) oral and general health-related risk behaviours. The employment of a follow-up survey at six months allowed for a longitudinal evaluation of behaviours in the above areas.

## Methodology

The study utilises a longitudinal, quantitative survey over 6-months.

### Recruitment

The recruitment process followed a convenience sampling approach, making use of advertisements such as the University research volunteering website, posters in university buildings, and university social media pages to attract participants. An initial sample size of 205 participants with comparisons between the two time points of 114 students. 205 participants volunteered and were recruited for the study and screening of participants was based on the eligibility inclusion criteria: (a) between 18-24 years old, (b) in their first year of university (c) studying or attending university for the first time, (d) living away from their family home, (e) access to a computer or mobile phone with internet connection, (f) able to comprehend and speak English, (g) available for the duration of the study and attending campus during the duration of the study and (h) complete an informed written consent form. The recruitment period started on 1^st^ October 2022 with recruitment end date on 31^st^ January 2023. The study design and protocol were approved by the University of Manchester Research Ethics Committee (Reference: 2023-13950-27733).

### Data collection

The study gathered data through online questionnaires using the online Qualtrics software. A customised questionnaire was designed, recognising the need to tailor it specifically for university students to maximise data collection. At baseline, a demographic questionnaire and a behaviour questionnaire were sent to participants. At follow-up, the behaviour questionnaire and feedback questionnaire were sent to participants. The demographic and behaviour questionnaire were developed and modified according to the public and participant involvement work completed prior to this study. All data collected was analysed descriptively (frequency, counts, trends) and assessed to explore changes over time. It was coded and analysed using SPSS (IBM SPSS Statistics for Macintosh, Version 25.0.). For dichotomous or nominal variables between groups, Chi-Squared test was used. To meet the requirement for the Chi-squared test, the individual expected count must be at least 1. Consequently, if any cell has an expected count below 1, sub-groups were merged to facilitate the Chi-squared test. Alternatively, if data grouping was not possible, the Fisher’s Exact test was used. For dichotomous variables within groups between time points (baseline and follow-up) McNemar test was conducted. For continuous variables between time points Wilcoxon Signed rank test was conducted. The level of statistical significance was set at p<0.05.

## Results

### Demographics of sampling characteristics

Excluding those who did not meet the inclusion criteria, there were 290 participants who consented. Of these 205 completed the study (70.7% response rate), with no dropouts at baseline. Participants were 66% female (n=135) and 34% male (n=70). Missing data appeared at random, with no discernible patterns. Mean age did not significantly differ between baseline and follow-up. Participants were ethnically diverse: White (52.2%), Asian or British Asian (32.25) and Black, Black British, Caribbean, or African (5.4%). At follow-up, 116 completed data collection (56.6% response rate); 89 dropped out from baseline. After excluding two with incorrect ID numbers, 114 completed data collection. Gender ratio: 68.4% female (n=78), 31.6% male (n=36). Ethnicity distribution was similar to baseline: White (46.5%), Asian or British Asian (32.5%), and Black, Black British, Caribbean, or African (7.0%).

### Demographic questionnaire including the use of technology

The attitudes towards technology and mobile device use were explored. The demographic data (found in the supplementary data) shows that students are engaging with technology and digital devices. The majority of students are on monthly mobile contracts (82.9%), and students are commonly using instant messaging platforms (54.1%), with the most common platforms being WhatsApp and Instagram Messenger (98.5% and 73.7%). Many students do not use a smartwatch or health device (68.3%), but there is a usage of fitness or health-related applications such as Apple Health and MyFitnessPal (34.7% and 18.4% respectively). Many students are also aware of the features of connected toothbrushes such as brush tracking technology, connection with an app, and use of mobile technology (94.6%).

### Oral health behaviours

**At baseline** oral health behaviours showed that many students consider their oral care routine, and the condition of the teeth and gums were average to good. Around half of students prioritise fresh teeth and clean teeth (58.0% and 49.1%) as being most important with regards to their mouths. Approximately 7 out of 10 students brush twice a day (70.2%) and around 3 in 10 students have experienced toothache in the duration of this study (32.5%). Just over half of students’ gums do not bleed when brushing teeth (60.5%). In relation to the dentist: the majority of responses indicated students visited the dentist less than 6 months ago (29.8%), for a routine check-up (63.2%) followed by treatment such as a filling or extraction (15.8). The majority have not registered with a dentist since starting university (95.6%) with 60.5% intending to continue visiting their current dentist. **At follow-up**, students now prioritise having white teeth and avoiding toothache (50.9% and 28.1%) as being most important with regards to their mouths. 68.4% of students are brushing their teeth twice a day and levels of bleeding gums is unchanged. In relation to the dentist: most students visited the dentist more than 1 year ago but less than 2 years ago for similar reasons to baseline. Students have continued to not register with a new dentist (95.6%) and more students intend to continue visiting their current dentist (78.1%). All data available in the S2 text.

### Risk behaviours

**Baseline** data on risk behaviours reveals that 10.5% of students smoke, mostly less than 5 cigarettes daily (83.3%), starting 6-12 months ago (50.0%). Vaping is less common, with 75.4% having never vaped and 9.6% currently doing so. Many students drink alcohol (67.5%) typically 2-4 times monthly or 2-4 times a week (both 32.5%), commonly consuming between 3-4 units or 5-6 units (both 28.6%) though consumption of 8 or more units varies. Most students exercise regularly, with 38.6% engaging more than once a week, primarily carrying out general fitness or at the gym (43.0%). Food intake is rated as average by 54.4%, with high consumption of sugary items like biscuits, cakes, cream cakes, and sweet pies (86.8%), often several times weekly (46.5%). Non-alcoholic drink intake is also believed to be average according to the participant responses from the Likert scale, with many consuming such drinks 1-2 times daily (87.5%), mostly sugary soft drinks (52.6%). Some students consume energy drinks (22.8%), with 88.5% reporting changes in consumption since starting university. **Follow-up** data on risk behaviours indicate that 7.0% of students smoke, with 75.0% smoking less than 5 cigarettes daily. Half of students who smoke stated they began smoking over a year ago but less than two years ago (50.0%). Regarding vaping, 11.4% of students vape, with 69.3% having never done so before. Many students drink alcohol (57.0%) monthly or less (43.1%) typically consuming 1-2 units (41.5%). The frequency of 8 or more units consumed varies but generally lower levels than baseline. Exercise frequency remains high, with 56.1% engaging at least once a week, often through activities like walking or going to the gym. From a Likert scale ranging from very unhealthy to very healthy, food intake is considered average or healthy (43.0% for both) with high consumption of sugary items (86.0%), often consumed several times weekly (45.6%). Non-alcoholic drink intake is also mostly believed by students to be average (40.4%) according to participant responses from the Likert scale, with many students consuming such drinks 1-2 times daily (68.4%), favouring sugary drinks like Cola (61.4%). About a third of students drink energy drinks, mostly 1-2 per day, with many reporting changes in drinking habits since starting university (94.4%). Baseline and follow-up comparisons were only made with students that completed both questionnaires at baseline and follow-up. All data available in the S3 text.

## Significant study data

There were significant associations between many variables in the study, including oral health behaviours and risk behaviours. At baseline, the associations that were seen are shown in Table 1 below and the associations at follow-up are shown in Table 2.

**Table 1:** Associations between different variables in the study behaviour data at baseline.

| Study data comparisons (baseline): | Direction of association | Test statistic<sup>a</sup>, P value<sup>b</sup>, Effect size<sup>c</sup> |
| --- | --- | --- |
| Exercise and oral care routine rating | Exercising more frequently = a better oral care routine rating | $X^2(8)=21.065$, $p=0.008$, Cramer's $V=0.227$ |
| Food intake and condition of the gums | Unhealthy food consumption = poorer self-reported condition of gums | $X^2(6)=16.786$, $p=0.010$, Cramer's $V=0.202$ |
| Frequency of food intake and condition of the teeth | Consuming unhealthy foods more frequently = less healthy teeth | $X^2(6)=22.414$, $p=0.001$, Cramer's $V=0.234$ |
| Non-alcoholic drink intake and oral care routine rating | Healthy non-alcoholic drink intake = better oral care routine rating | $X^2(6)=16.890$, $p=0.010$, Cramer's $V=0.203$ |

<sup>a</sup>Chi-squared test with degrees of freedom. <sup>b</sup>Significance level is set at $p<0.05$ . <sup>c</sup>Cramer's V value.

**Table 2:**
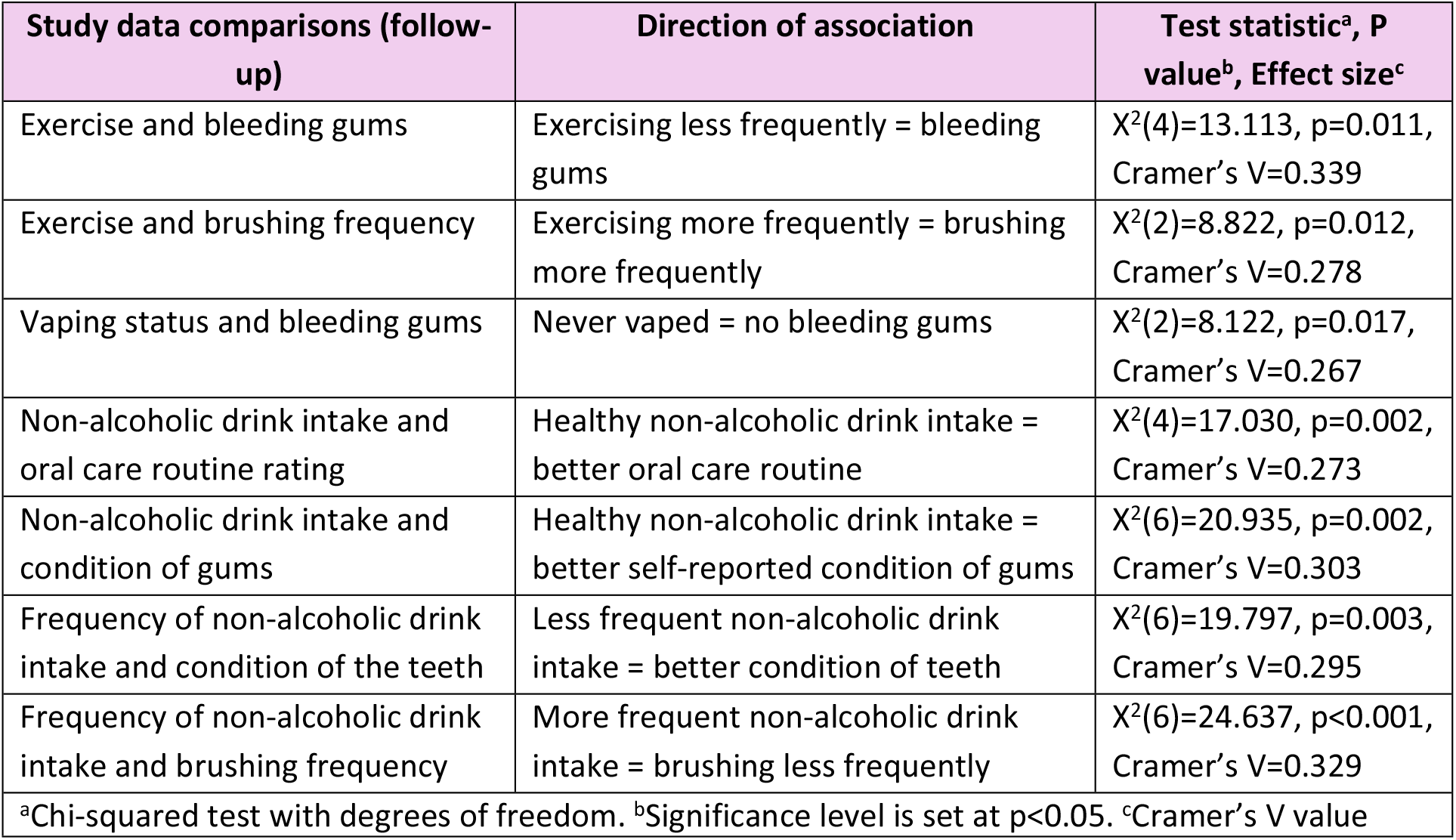
Associations between different variables in the study behaviour data at follow-up.

## Comparisons between baseline and follow-up

Associations between variables in the study data at the two time-points (baseline and follow-up) are shown below in Table 3.

**Table 3:** Associations between variables in the study data between the two time-points.

| Comparisons significances over time points: | Test statistic<sup>a</sup>, P value<sup>b</sup>, Effect size<sup>c</sup> |
| --- | --- |
| Condition of the teeth ↓ | Z=-3.499, p<0.001, r=-0.32 |
| Condition of the gums ↓ | Z=-2.873, p=0.004, r=-0.27 |
| Brushing frequency ↓ | Z=-2.987, p=0.003, r=-0.27 |
| How long since last visit to the dentist ↑ | Z=-2.497, p=0.013, r=-0.23 |
| Intend to visit home dentist ↑ or new dentist ↓ | Z=-3.035, p=0.002, r=-0.28 |
| Alcohol consumption frequency ↓ | Z=-2.169, p=0.030, r=-0.20 |
| How many units of alcohol ↓ | Z=-5.651, p<0.001, r=-0.53 |
| 8 or more units of alcohol ↓ | Z=-2.450, p=0.014, r=-0.23 |
| Frequency of food intake ↓ | Z=-2.903, p=0.004, r=-0.27 |
| Non-alcoholic drink intake ↓ | Z=-2.083, p=0.037, r=-0.20 |
| <sup>a</sup>Wilcoxon signed-rank test, <sup>b</sup>Significance level is set at p<0.05, <sup>c</sup>Correlation coefficient r statistic, ↑ refers to an increase over study period, ↓ refers to a decrease over study period |  |

Overall, many patterns and trends were seen in the data that show differences in time in the oral health behaviours and risk behaviours that were explored in this demographic. Some oral health behaviours saw no statistical differences over time such as the rating of oral care routine (p=0.075), bleeding gums (p=0.567) and toothache (p=0.410). Some risk behaviours saw no statistically significant differences over time such as smoking (p=0.401), vaping (p=0.431), happiness regarding weight (p=0.488), energy drink consumption (p=0.253), energy drink frequency (p=0.194), changes in energy drinks (p=0.105). Changes over time were also not seen in exercise levels (p=0.976). In relation to dental attendance, the importance of attending the dentist regularly saw no statistical difference over time (p=0.324), as well as a reason to visit the dentist (p=0.687). More detailed non-significant comparisons can be seen in the S1 text. In summary, oral health behaviours, outcomes associated with risk behaviours, and risk behaviours over 6 months were explored. Poor oral health behaviours are associated with some risk behaviours and some behaviours have not changed since starting university.

## Discussion

### Summary of findings

The data can be used to evaluate the oral health behaviours and risk behaviours of university students. The study reveals a comprehensive association between various lifestyle and oral health outcomes among university students. Specifically, alcohol consumption, vaping, and regular consumption of unhealthy drinks are all correlated with oral health such as deteriorating condition of the teeth, rating of oral care routine, and bleeding gums. Infrequent dental visits are prevalent among most university students. However, overweight status does not exhibit a significant association with these variables, while exercise remains widespread within this demographic. Importantly, the study also highlights an overall reduction in brushing frequency among university students.

The findings from the demographic analysis reveal several key trends among the student population surveyed such as regular usage of text messages and instant messaging platforms among students, and the vast majority of students are aware of the functionalities offered by connected toothbrushes, with 80.5% recognising their connection capabilities with a mobile application. These findings strongly suggest that the demographic under study has a widespread reliance on technology for various purposes, including communication, fitness tracking, and health management. This reflects the use of technology by university students and young adults [20].

The data revealed at baseline, students’ top priority was having ‘fresh teeth’ or ‘clean teeth’ while at follow-up, their priorities or concerns shifted to having ‘white teeth.’ This suggests that students have shifted their priorities. Additionally, concerns about appearance among this age group may have prompted students to prioritise having white teeth. Current research supports these findings, as the appearance of teeth impacts an individual’s attractiveness and plays a role in social interactions [21, 22]. Given that young adults are often more conscious of their appearance, especially in social settings [23], may cause individuals to have the desire to have white teeth [24]. This underscores the importance of social interactions during a student’s time at university [25] and the importance of dental appearances in shaping perceptions and interactions [26].

The study findings raise awareness of the lack of registration for new dental services and most students intending to visit their previous dentist and not find a new dentist whilst at university. This may be due to the lack of new NHS dentists taking on new patients during these times due to capacity [27]. Also, the last visits to the dentist have gone from ‘less than 6 months ago’ to ‘more than 1 year ago but less than 2 years ago.’ Suggesting, most students have not attended the dentist whilst at university at the time points. More research needs to be explored to assess whether students do visit the dentist whilst at university with normal university courses being between 3-5 years and an NHS dentist registration period of only 2 years [28, 29], this may lead to many students not having an NHS dentist upon finishing university.

The rating of oral care routines is influenced by numerous factors, as indicated by their correlations with exercise, and non-alcoholic drink intake. These associations show the intertwined nature of oral health behaviours with broader lifestyle choices. Research supports the notion that regular exercise can safeguard against diseases and promote overall well-being [30], including oral health benefits such as preventing periodontal disease [31]. This study also assessed links between exercise and brushing frequency, showing that students who are more physically active are more likely to brush twice a day, current research also supports this notion [32]. Moreover, a deterioration in students’ oral health over time is evident, with the worsening of self-reported conditions of the teeth and gums. The prevalence of bleeding gums among university students aligns with existing research findings [33]. There is also an association between bleeding gums and vaping gums, indicating potential long-term implications of this behaviour on gum health, with existing research also demonstrating this link [34, 35, 36].

The study findings showed that there has been a decrease in students who are brushing twice a day and an increase in students who are brushing once a day (Z=-2.987, p=0.003). This supports the notion that many students only brush once a day, as highlighted by Bashiru and Anthony 2014, who found that 90% of students brushed their teeth once a day. The reasons for this decrease in brushing frequency may be due to busy schedules, lack of routines, late nights studying or out partying during their time at university. It could be speculated that factors such as increased workload, stress, or changes in routine such as late nights may have contributed to this trend. There is a correlation between brushing frequency, neglecting toothbrushing and stress [37].

In this study, unhealthy food and drinks are associated with the condition of the gums, and it has been shown that dietary habits may detrimentally impact oral health, leading to increased risks of gum disease [38, 39]. The frequency of unhealthy dietary food and drink consumption has shown links with the condition of the teeth too, indicating that poor dietary habits negatively impact oral health outcomes. The decline in the consumption frequency of unhealthy foods indicates a positive shift towards a more balanced dietary pattern over time. This transition is important as it suggests potential benefits for dental health by reducing the risk of tooth decay and dental issues [40]. By decreasing the frequency of consuming sugary and unhealthy foods, students can minimise the exposure of their teeth to sugars and acids from the food, thereby reducing the likelihood of enamel demineralisation, tooth weakening, erosion, and cavities [41]. Therefore, healthy dietary habits are crucial to prevent tooth decay [42] and by cutting back on consumption, students can improve their oral health.

Alcohol consumption over time decreased overall, with reduced consumption, reduced units of consumption and fewer students binge drinking. The existing evidence supports this notion as students drink more alcohol at the start of university [43]. The trends in non-alcoholic drink intake among students reveal a prevalent consumption of sugary beverages, despite many considering their intake as average. This pattern suggests a normalisation of sugary drink consumption among students.

Moreover, the findings imply a connection between non-alcoholic drink intake and, the condition of the teeth and oral care routine. Individuals who consume unhealthy foods may consume more sugary drinks, thereby raising concerns about potential implications for dental health such as tooth decay and erosion [44, 45]. In addition, 31.6% of students reported consuming energy drinks which is an increase over time, with 94.4% of them indicating a change in consumption since starting university, aligning with previous research on energy drink consumption among students. Possible motivations for this trend include improving academic performance, combating fatigue, socialising, and enjoying leisure time with friends [46].

## Key results and links

Data analysis revealed changes over time: (a) self-reported deterioration of the condition of teeth and gums over time, (b) students believe the avoidance of toothache and having white teeth are most important at follow-up, compared to fresh and clean teeth at baseline, (c) reduced brushing frequency over time, (d) consistent levels of toothache at two time points but a general prevalence of toothache overall, (e) less frequent visits to the dentist, (f) check-ups and treatments (such as fillings and/or extractions) are the most common reason for visits to the dentist, (g) students not registering with new dentists, (h) general importance of attending the dentist visible, (i) increased in students intending to visit their previous dentists, (j) general high levels of alcohol consumption, (k) decrease in alcohol consumption frequency over time, (l) decreased in the number of units of alcohol consumed over time, (m) decrease over time in the number of students consuming 8 or more units of alcohol on one occasion (binge drinking), (n) general belief that smoking, alcohol and vaping effects the mouth, (o) consensus that weight is average, (p) consistently high levels of exercise over time, (q) unhealthier food intake at baseline but reduction in consumption of unhealthy foods over time, (r) unhealthy non-alcoholic drink intake but improvements over time, (s) consistently high levels of energy drink consumption over time, (t) increase in energy drink consumption since starting university.

## Direction of future work

Future research could focus on developing interventions specifically designed for university students aimed at addressing improving attitudes, oral health behaviours and associated risk behaviours prevalent during their academic period.

## Strengths

Feedback from participants, collected through the end-of-study questionnaire, reflects high levels of satisfaction and enjoyment, with 91.1% expressing enjoyment in participation and 96.5% agreeing on the importance of the study topic. These results underscore strong support for the study. The preference for the online questionnaire format suggests the demographics’ comfort and familiarity with digital platforms, reflecting their frequent engagement with online platforms and the potential to use digital technology for future research endeavours or interventions. The study participants also studied eight of the most enrolled-in degree programmes offered at the University of Manchester [47]. The use of a diverse sample with students enrolled in a wide range of academic subjects constitutes a strength of this study. By including participants from various fields of study, the research encompassed a broad spectrum of perceptive, enhancing the richness and representativeness of the data collected. Importantly, this approach facilitated the capture of insights from individuals not exclusively studying health-related subjects, who may be more health conscious.

## Limitations

The reliance on self-reported data for assessing behaviours and attitudes introduces potential limitations due to social desirability and recall biases, as participants may provide responses that conform to societal norms rather than accurately reflecting their behaviours. Efforts were made to mitigate this bias through confidentiality assurances. The anonymity provided by online questionnaires may have eliminated potential social pressures or biases that could arise in face-to-face interactions. Therefore, leveraging digital platforms for data collection not only maximises participant engagement but also ensures a more efficient and inclusive process. Additionally, the dropout of participants from baseline to 6 months resulted in a smaller sample size than expected, meaning that data from participants who only completed the baseline questionnaire could not be included in the analysis of the comparisons between the two time periods. This attrition could have introduced potential attrition bias; however, the characteristics of the participants who completed the study showed no apparent difference from those who dropped out.

## Conclusion

The findings provide a comprehensive insight into the complex network of behaviours and attitudes among university students, revealing the prevalence of detrimental oral health behaviours and associated risk behaviours. Health outcomes and behaviours such as oral care routine rating, condition of the teeth, condition of the gums, bleeding gums, brushing frequency, exercise, vaping, and unhealthy dietary habits are interconnected, indicating a complex interplay between various lifestyle choices. The study emphasises the critical period of university life where unhealthy behaviours often emerge and potentially solidify into lifelong habits. Finding a new dentist and the lack of visits to the dentist during their time at university is concerning, highlighting the imperative for further initiatives aimed at promoting dental attendance among university students while they are residing away from home.

## Data Availability

All relevant data are within the manuscript and its Supporting Information files.

## Acknowledgments

The authors would like to thank all participating students involved in this study.

